# Cryptic Transmission of the Delta Variant AY.3 Sublineage of SARS-CoV-2 among Fully Vaccinated Patients on an Inpatient Ward

**DOI:** 10.1101/2021.08.05.21261562

**Authors:** Katherine Linsenmeyer, Kalpana Gupta, Rebecca Madjarov, Michael E. Charness

## Abstract

**Background:** Recent reports indicate that vaccination is effective in reducing symptomatic infection with the Delta variant of SARS-CoV-2 (DV) but is less protective against asymptomatic transmission of DV in outpatients than for earlier variants.

Here we report cryptic transmission associated with high DV viral load among vaccinated patients on an inpatient medical-surgical ward.

**Methods:** This observational study included all persons diagnosed with breakthrough SARS-CoV-2 infections at the VA Boston Healthcare System (VABHS) from March 11, 2021 to July 31, 2021, including those tested for surveillance, admission, symptoms, and as part of an outbreak investigation in July 2021. SARS-CoV-2 infection was diagnosed by reverse-transcription polymerase chain reaction (PCR) (Cepheid). Variants were identified by MassARRAY SARS-CoV-2 Variant Panel (36-plex PCR, Agena BioScience) for most breakthrough cases after June 2021 Viral genomic sequencing was performed by the Jackson Laboratory.

**Results:** An inpatient was diagnosed with asymptomatic DV infection on routine pre-discharge testing. Contact tracing detected infection in 6 of 38 patients (15.8%), 1 of 168 staff (0.6%), and 1 of 6 visitors (16.7%). Infection at the time of diagnosis was asymptomatic in 4 proximate, vaccinated patients, 1 vaccinated visitor, and 1 vaccinated employee caring for 1 undiagnosed, infected, vaccinated patient. Patients were unmasked, whereas staff wore surgical masks. PCR cycle threshold (Ct) for breakthrough infections indicated more than 1000-fold higher viral load for DV (Ct:21.7±4.3; n=15) than for earlier variants (Ct: 31.8±10.9, n=12; p=.003 (*t*-test)).

**Conclusion:** These findings demonstrate transmission of DV with high viral load between vaccinated inpatients, the continued efficacy of masking and vaccination for protecting healthcare personnel, and the potential need for post-admission surveillance to prevent cryptic DV transmission.

## Introduction

The course of the COVID-19 pandemic has been punctuated by the emergence of increasingly transmissible SARS-CoV-2 variants and the development of efficacious vaccines. Risk mitigation measures in community and hospital settings have evolved in parallel to align protective measures with changing levels of risk. Vaccines have proven highly efficacious in preventing severe disease and death due to a variety of SARS-CoV-2 variants.^1-5^ Vaccination also reduces viral load, asymptomatic infection, and viral transmission for many variants.^6-9^

Since March of 2021, the highly transmissible Delta variant has become predominant in many countries across the globe. Recent reports indicate that in contrast to earlier variants, the Delta variant achieves equally high viral loads in the nasopharynx of vaccinated and unvaccinated persons.^10-11^ Furthermore, transmission in a community setting may occur between vaccinated persons.^11^ Here we demonstrate that even in the highly controlled environment of an inpatient medical-surgical ward, cryptic transmission of the Delta variant may occur among vaccinated patients.

## Methods

The study cohort included all persons with breakthrough SARS-CoV-2 infections at the VA Boston Healthcare System (VABHS) from March 11, 2021 to July 31, 2021. Testing was conducted for surveillance, admission, symptoms, and as part of an outbreak investigation in July 2021. Breakthrough infections were defined as those for which likely exposure occurred more than two weeks after completion of a full vaccination series. Most vaccinated patients and healthcare workers (HCW) received mRNA-1273 (Moderna) vaccine. At the end of the study period, 91% of HCW and 55% of patients were fully vaccinated. In the inpatient setting, HCW wore surgical masks and patients did not.

SARS-CoV-2 infection was diagnosed by reverse-transcription polymerase chain reaction (PCR) (Cepheid),^12^ and cycle threshold (Ct) was recorded for all HCW and patients with breakthrough infections during the study period. A difference of 3 amplification cycles was estimated by the manufacturer to represent a 10-fold difference in viral load. Variants were identified by MassARRAY SARS-CoV-2 Variant Panel (36-plex PCR, Agena BioScience) for the majority of breakthrough cases after June 2021. During July of 2021, 9 of 10 samples from VABHS and 27 of 28 from VA New England Healthcare System were identified as the Delta variant. Sequencing was performed by the Jackson Laboratory. For samples that were not typed by PCR or sequencing, all SARS-CoV-2 infections arising after June 10 were presumed to be due to the Delta variant, and all infections arising before June 10 were presumed to be due to other variants.^11,13^ Between March and June 10, the predominant variants in New England were Alpha, Gamma, and D614G.^13^

Infections in the outbreak described below were diagnosed between July 16 and July 26, 2021. Following the recognition of the first hospital cases, surveillance of exposed HCW was carried out using daily antigen testing (Abbott BinaxNow) for indirect contacts and PCR for direct contacts or confirmation of positive results. PCR was employed for testing of all exposed patients.

Data are expressed as mean ± standard deviation, and statistical difference between means was evaluated with the Students *t-*test or *X*^*2*^. Statistical significance was defined as p < .05 using two-tailed tests.

The VABHS Institutional Review Board determined that this was a quality improvement project and exempted this study from review.

## Results

An outbreak of the Delta variant of SARS-CoV-2 on an inpatient ward was discovered during routine pre-discharge testing of an asymptomatic, vaccinated patient (Patient F in Figure 1A) and subsequent contact tracing of 38 patients, 168 HCW, and 6 visitors. This extensive testing of HCW revealed just one infected, vaccinated HCW (F1) who worked in close contact with Patient F.

**Figure 1.**
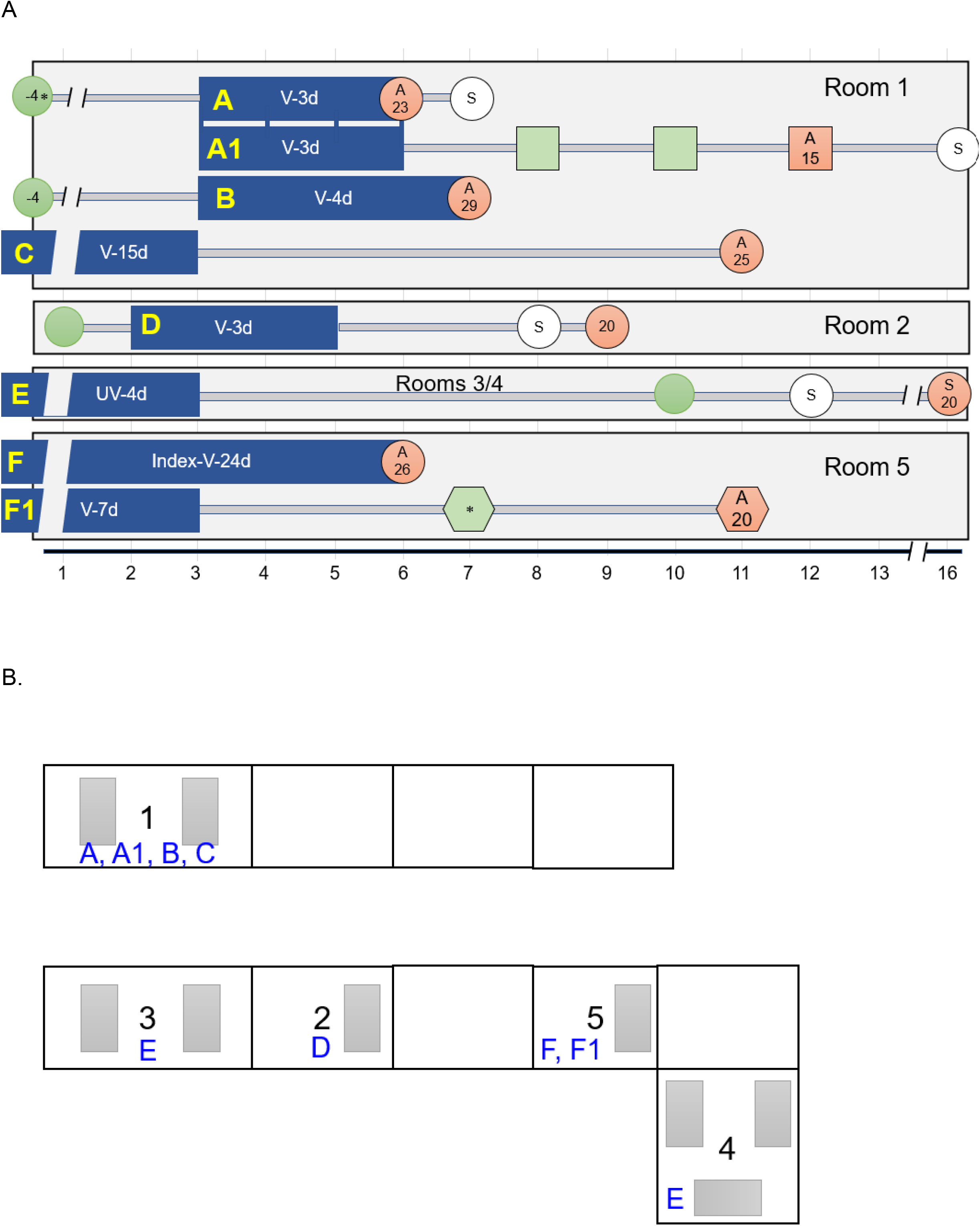
Evolution of SARS-CoV-2 infections distributed across time for persons with exposures in a single inpatient unit. A. Number scale at the bottom of the figure indicates sequential days with indicated breaks in the scale. Blue bars represent days spent in the indicated rooms on a single medical-surgical unit. Circles represent patients; square, a family member; hexagon, a healthcare worker. Numbers within shapes indicate PCR Ct. Red fill indicates positive PCR test; green fill, negative PCR; * within green circle indicates a negative antigen test. Negative numbers in circles represent days prior to the beginning of the timeline. V, vaccinated; UV, unvaccinated; Nd appearing after V- or UV- represents number of days on the outbreak unit. A, asymptomatic at time of diagnosis; S, symptomatic at time of diagnosis. Open circles indicate the onset of symptoms prior to diagnosis. The first diagnosed patient is designated as Index. B. Location of rooms. Letters correspond to rooms identified in Figure 1A. Grey rectangles represent beds and together depict 1-bed, 2-bed, and 3-bed rooms.

The first 5 of 6 infected patients (Patients A-D and F) diagnosed with infection were fully vaccinated, as were a visiting family member and the infected HCW (Figure 1A). Patient E was unvaccinated. Four of the first 5 patients (Patients A,B,C,F) were asymptomatic at the time of diagnosis. Two infected patients (A,B) shared a room vacated two hours earlier by another patient (C) who was asymptomatic upon discharge and PCR-positive 8 days later. The remainder of the infected patients (D-F) occupied nearby rooms (Figure 1B). An infected, vaccinated patient with dementia (Patient F) wandered into several of these rooms prior to diagnosis. An infected, vaccinated patient (A) likely infected a visiting family member (A1), who had two negative PCR tests (days 1 and 3) followed by a positive test on day 6 (Ct = 15) following their last exposure. One vaccinated patient (A) developed hypoxemic respiratory failure and improved following intensive care, the family member (A1) required hospitalization, and the unvaccinated patient (E) required intensive care and died.

Samples were submitted for variant typing on 7 of the 8 persons in the outbreak, and all 7 samples were positive for the Delta variant; genomic sequencing on 4 of 4 indicated the Delta variant sublineage AY.3. PCR Ct in all breakthrough infections was significantly lower for the Delta variant (21.7±4.3; n=15) than for earlier variants (31.8±10.9, n=12; p=.003 (*t*-test))(Figure 2), suggesting a more than 1000-fold higher viral load for the Delta variant compared to earlier variants. The two groups (Delta vs earlier variants) did not differ in age (68.0±18.8 vs. 53.3±26.2; p = .10), sex (male) (73.3% vs 66.7%; p = .71), or occurrence of symptoms at the time of diagnosis (33.3% vs 50%; p = .38).

**Figure 2.**
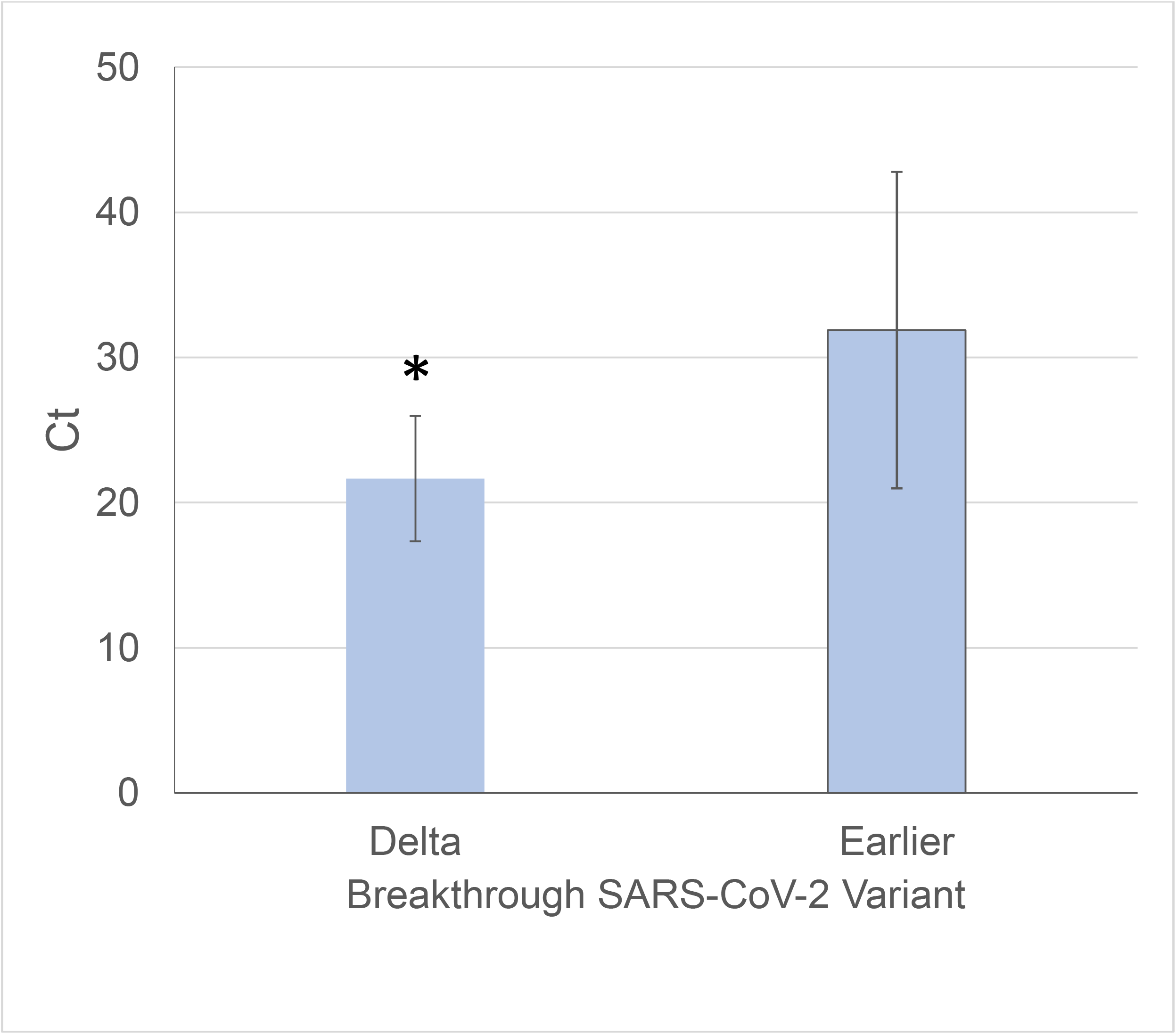
PCR cycle threshold (Ct) in breakthrough infections with the Delta variant vs earlier variants of SARS-CoV-2. Shown are the mean ± standard deviation for the two groups. * p = .003.

## Discussion

Cryptic transmission of the Delta variant occurred among asymptomatic, vaccinated inpatients on a single inpatient ward. The source may have been a patient whose infection arose after preadmission testing from a pretesting exposure or from a subsequent hospital exposure that was not identified despite extensive contact tracing. Infection spread among vaccinated patients and extended to a vaccinated HCW and a vaccinated visiting family member. This cluster might not have been discovered save for a routine discharge test that triggered follow-up contact tracing, because most patients were asymptomatic when diagnosed, and some patients were discharged prior to the discovery of the outbreak. These observations contrast with a report that unvaccinated persons were the sole source for breakthrough infections in HCW with the Alpha variant.^8^

High viral load increases the risk for transmission of SARS-CoV-2;^14^ therefore, the high viral load of the Delta variant in asymptomatic vaccinated persons likely increases the risk for nosocomial transmission. Because the incubation period for SARS-CoV-2 infection is 2-14 days, screening for SARS-COV-2 infection, even on the day of admission, will not detect any infections arising from exposure on the day prior to admission or many infections arising from earlier exposures. Hence, the use of sensitive tests^15^ for postadmission surveillance of inpatients, regardless of vaccination status, may be necessary to prevent nosocomial infection during surges of highly transmissible variants.

Though reports have identified transmission from HCW to patients as an important route for nosocomial SARS-CoV-2 infection,^16,17^ here, transmission likely occurred between patients and from patients to the sole infected HCW and to a visitor. Of note, infection spared all but one of 168 HCW that were exposed to infected patients. That transmission was primarily among patients and not between patients and HCW perhaps reflected the fact that HCW wore surgical masks and patients did not. Hence, despite the high viral load of their patients, masking combined with vaccination may have protected HCW during this outbreak. Encouragement of patient masking might further decrease nosocomial Delta variant infection.

Our findings support a growing literature that vaccination does not prevent the development of high viral load in the nasopharynx of persons infected with the Delta variant or the propensity for asymptomatic transmission,^10,11^ in contrast to findings with earlier variants.^6-9^ Brown et. al. ^11^ recently described community transmission of the Delta variant B.1.617.2 in a cohort of predominantly vaccinated persons who congregated unmasked in indoor spaces. High viral load did not differ between vaccinated and unvaccinated persons. In that study, sequencing identified the Delta variant (B.1.617.2) in 89% of cases and the Delta AY.3 sublineage in just 1%. In the current study, sequencing identified the Delta AY.3 sublineage 4 of 4 cases. Our observations indicate that transmission of the Delta variant sub-lineage AY.3 may occur among vaccinated persons even in the more controlled environment of an inpatient medical-surgical ward. It remains to be determined whether the Delta variant AY.3 lineage is more transmissible than the Delta Variant B.1.617.2. Our findings do raise concern that without stricter risk mitigation, nosocomial transmission of the Delta variant and Delta sublineages will occur more frequently than with prior, less transmissible variants, even in vaccinated persons.

This observational study has several limitations. The outbreak included a small number of individuals. The absence of full viral genome sequencing on all 8 cases prevented clear identification of the source and trajectory of viral transmission. One case was not typed to positively identify the Delta variant. Finally, though the data demonstrate that asymptomatic infection and transmission of the Delta variant may occur in vaccinated persons, this study does not address the relative risk for Delta variant infection in vaccinated versus unvaccinated persons.

## Data Availability

Aggregate data files are available after approval by institutional IRB and Privacy Officer.

## Acknowledgements

We are grateful to Gary Stack, M.D., Ph.D., for analysis and interpretation of the PCR variant typing data, Michael A. Gelman, M.D., Ph.D., for thoughtful analysis and discussion of the outbreak, Stephen Brecher, Ph.D., for assistance with molecular testing, Judy Strymish, M.D., for clinical expertise, and VABHS Infection Prevention, Nursing Service, Occupational Health, and Testing Teams for excellent support in limiting this outbreak.

## References

1. Baden LR, El Sahly HM, Essink B, et al. Efficacy and Safety of the mRNA-1273 SARS-CoV-2 Vaccine. N Engl J Med. 2020 Dec 30. doi:10.1056/NEJMoa2035389

2. Polack FP, Thomas SJ, Kitchin N, et al. Safety and Efficacy of the BNT162b2 mRNA Covid-19 Vaccine. N Engl J Med. 2020 Dec 31;383(27):2603–2615. doi: 10.1056/NEJMoa2034577.Epub2020Dec10

3. Barouch DH, Stephenson KE, Sadoff J. Durable Humoral and Cellular Immune Responses 8 Months after Ad26.COV2.S Vaccination. N Engl J Med 2021

4. Lopez Bernal J, Andrews N, Gower C, et al. Effectiveness of Covid-19 vaccines against the B.1.617.2 (delta) variant. N Engl J Med. DOI: 10.1056/NEJMoa2108891l

5. Nasreen S, He S, Chung H, et al. Effectiveness of COVID-19 vaccines against variants of concern, Canada. medRxiv 2021.06.28.21259420; doi: https://doi.org/10.1101/2021.06.28.21259420

6. Dagan N, Barda N, Kepten E, et al. BNT162b2 mRNA Covid-19 Vaccine in a Nationwide Mass Vaccination Setting. N Engl J Med. 2021 Apr 15;384(15):1412–1423. doi: 10.1056/NEJMoa2101765. Epub 2021 Feb 24. PMID: 33626250; PMCID: PMC7944975.

7. Thompson MG, Burgess JL, Naleway AL, et al. Prevention and Attenuation of Covid-19 with the BNT162b2 and mRNA-1273 Vaccines. N Engl J Med. 2021 Jul 22;385(4):320–329. doi: 10.1056/NEJMoa2107058. Epub 2021 Jun 30. PMID: 34192428; PMCID: PMC8262622.

8. Bergwerk M, Gonen T, Lustig Y, et al. Covid-19 Breakthrough Infections in Vaccinated Health Care Workers. N Engl J Med. 2021 Jul 28. doi: 10.1056/NEJMoa2109072. Epub ahead of print. PMID: 34320281.

9. Gohil SK, Huang SS. Community COVID-19 Incidence and Health Care Personnel COVID-19 Seroprevalence. JAMA Netw Open. 2021 Mar 1;4(3):e211575. doi: 10.1001/jamanetworkopen.2021.1575. PMID: 33688960.

10. Li B, Deng A, Li K, et a. Viral infection and transmission in a large, well-traced outbreak caused by the SARS-CoV-2 Delta variant medRxiv 2021.07.07.21260122; doi: https://doi.org/10.1101/2021.07.07.21260122

11. Brown CM, Vostok J, Johnson H, et al. Outbreak of SARS-CoV-2 Infections, Including COVID-19 Vaccine Breakthrough Infections, Associated with Large Public Gatherings - Barnstable County, Massachusetts, July 2021. Morb Mortal Wkly Rep. ePub:30 July 2021.DOI: http://dx.doi.org/10.15585/mmwr.mm7031e2

12. Gupta K, O’Brien WJ, Bellino P, et al. Incidence of SARS-CoV-2 infection in healthcare workers after a single dose of mRNA-1273 vaccine. JAMA Network Open, 2021 PMID: 34132795

13. CDC. SARS-CoV-2 Variant Classifications and Definitions. https://www.cdc.gov/coronavirus/2019-ncov/variants/variant-info.html, accessed July 31, 2021

14. Marks M, Millat-Martinez P, Ouchi D, et al. Transmission of COVID-19 in 282 clusters in Catalonia, Spain: a cohort study. Lancet Infect Dis. 2021 May;21(5):629–636. doi: 10.1016/S1473-3099(20)30985-3. Epub 2021 Feb 2. Erratum in: Lancet Infect Dis. 2021 Jul 5;: PMID: 33545090; PMCID: PMC7906723.

15. Frediani, J.K., Levy, J.M., Rao, A. et al.. Multidisciplinary assessment of the Abbott BinaxNOW SARS-CoV-2 point-of-care antigen test in the context of emerging viral variants and self-administration. Sci Rep 11, 14604 (2021). https://doi.org/10.1038/s41598-021-94055-1

16. Lucey M, Macori G, Mullane N, et al. Whole-genome Sequencing to Track Severe Acute Respiratory Syndrome Coronavirus 2 (SARS-CoV-2) Transmission in Nosocomial Outbreaks. Clin Infect Dis. 2021 Jun 1;72(11):e727–e735. doi: 10.1093/cid/ciaa1433. PMID: 32954414; PMCID: PMC7543366.

17. Borges V, Isidro J, Macedo F, et al. Nosocomial Outbreak of SARS-CoV-2 in a “Non-COVID-19” Hospital Ward: Virus Genome Sequencing as a Key Tool to Understand Cryptic Transmission. Viruses. 2021 Apr 1;13(4):604. doi: 10.3390/v13040604. PMID: 33916205; PMCID: PMC8065743.

